# Evaluating the impacts of tiered restrictions introduced in England, during October and December 2020 on COVID-19 cases: A synthetic control study

**DOI:** 10.1101/2021.03.09.21253165

**Authors:** Xingna Zhang, Gwilym Owen, Mark Green, Iain Buchan, Ben Barr

## Abstract

**Background:** In 2020, a second wave of COVID-19 cases unevenly affected places in England leading to the introduction of a tiered system of controls with different geographical areas subject to different levels of restrictions. Whilst previous research has examined the impact of national lockdowns on transmission, there has been limited research examining the marginal effect of differences in localised restrictions or how these effects vary between socioeconomic contexts. We therefore examined how Tier 3 restrictions in England implemented between October-December 2020, which included additional restrictions on the hospitality sector and people meeting outdoors affected COVID-19 case rates, compared to Tier 2 restrictions, and how these effects varied by level of deprivation.

**Methods:** We used data on weekly reported COVID-19 cases for 7201 neighbourhoods in England and adjusted these for changing case-detection rates to provide an estimate of weekly SARS-CoV-2 infections in each neighbourhood. We identified those areas that entered Tier 3 restrictions at two time points in October and December, and constructed a synthetic control group of similar places that had entered Tier 2 restrictions, using calibration weights to match them on a wide range of covariates that may influence transmission. We then compared the change in weekly infections between those entering Tier 3 to the synthetic control group to estimate the proportional reduction of cases resulting from Tier 3 restrictions compared to Tier 2 restrictions, over a 4-week period. We further used interaction analysis to estimate whether this effect differed based on the level of socioeconomic deprivation in each neighbourhood and whether effects were modified by the prevalence of a new more infectious variant of SARS-CoV-2 (B.1.1.7) in each area.

**Results:** The introduction of Tier 3 restrictions in October and December was associated with a 14% (95% CI 10% to 19%) and 20% (95% CI 13% to 29%) reduction in infections respectively, compared to the rates expected if only Tier 2 restrictions had been in place in those areas. We found that effects were similar across levels of deprivation and limited evidence that Tier 3 restrictions had a greater effect in areas where the new more infectious variant was more prevalent.

**Interpretation:** Additional restrictions on hospitality and meeting outdoors introduced in Tier 3 areas in England had a moderate effect on transmission and these restrictions did not appear to increase inequalities, having a similar impact across areas with differing levels of socioeconomic deprivation. Where transmission risks vary between geographical areas a tiered approach of local restrictions on outdoor mixing and hospitality can contribute to control of SARS-CoV-2 and is unlikely to increases inequalities in transmission.

## Introduction

In the latter half of 2020, England experienced a second wave of COVID-19 cases with prevalence increasing 10-fold from 0.1% in August to 1% in October.^1^ Reported cases of COVID-19 were unevenly distributed across the country, with areas in the North of England most severely affected. Initially a variety of local restrictions were introduced, then in October a standardised three Tier system was introduced. This was followed by a month-long national lockdown, after which tiered restrictions were re-introduced at the beginning of December, before a further national lock-down at the beginning of January 2021.

During the pandemic, evidence has accumulated showing that restrictions such as closing schools, bans on public events, bans on public gatherings, requirements to stay at home, and limits on internal movement have had substantially reduced transmission of SARS-CoV-2.^2^ Most of this evidence is, however, based on national implementation of policies^2^ and cross-country comparisons,^2^ that may not be applicable to restrictions that vary across relatively small areas within countries. Much of the evidence is also based on simulation studies, rather than empirical evidence, whereby observed changes in mobility and survey-based indicators of social contact following interventions are used to predict the expected impact of restrictions on cases and hospitalisations using compartmental models.^3^ Estimates of intervention impact from such studies may differ from the observed impact on cases, if the actual relationships between mobility and transmission following intervention differ from that assumed in the simulation. There is limited evidence comparing more vs less stringent localised restrictions within the same country and empirically estimating the impact of a marginal differences in stringency.

There are also concerns that such restrictions may have differential effects between different socioeconomic contexts. This is because people in more disadvantaged communities may not, for example, be able to comply with requirements to work from home due to their occupation or communications of the need to comply with restrictions, which have not been appropriately adapted to their target audiences, may be less effective.^4^ Additionally some groups may be less inclined to comply with restrictions due to mistrust of authorities. Understanding the potential differences in effect of control measures by socioeconomic group is important because the impact of the pandemic has disproportionally affected more disadvantaged groups.^5^ Interventions designed to control the pandemic may further exacerbate these inequalities if we do not evaluate their differential effects and take actions to mitigate any intervention-generated inequalities, or ideally prioritise approaches that reduce inequalities.^6^ Despite these warnings, there has been little investigation of the differential impact of such control measures between different socioeconomic contexts.

In this study we analyse the impact on SARS-CoV-2 transmission of Tier 3 restrictions introduced in October and December in England, compared to Tier 2 restrictions, where the main differences were additional restrictions on meeting people outdoors and restrictions on the hospitality sector in Tier 3 areas. We further investigate whether these effects varied between small areas based on their level of deprivation.

### Research in context

#### Evidence before this study

We searched Google Scholar, PubMed, bioRxiv, and medRxivfor English-language articles with the search terms (“COVID-19” OR “SARS-CoV-2” OR “coronavirus”) AND (“tier”) AND (“evaluation” OR “natural experiment” OR “difference in differences”). We found a large number of studies which explore the effect of non-pharmaceutical interventions on transmission of COVID-19. Most of these used mathematical models to simulate the effects of these interventions rather than empirical evaluations. A few studies exploited variations in restrictions between countries to empirically estimate effects of specific interventions.^7–13^ These studies provided evidence that restrictions such as closing educational institutions, restricting large gatherings and closing face to face businesses have been effective at reducing transmission. We identified only two studies investigating the impact of local restrictions. One simulation study predicted, based on contact and mobility data, that moving into Tier 3 restrictions in England in October reduced the effective reproduction rate by 10%.^3^ Another study used a difference-in-difference approach and found ambiguous effects depending on the time lags used.^14^ Difference-in-difference methods are however likely to give biased estimates with infectious disease outcomes when the comparison groups have differences in prevalence at baseline, as the exponential growth in both groups in the absence of intervention will not meet the parallel trends assumption.

### Added value of this study

Our study adds to the literature in a number of ways. Firstly, to our knowledge, this is the first study to empirically estimate the impact on SARS-CoV-2 infections of the additional restrictions applied to Tier 3 areas in England compared with the counterfactual of having only applied Tier 2 restrictions. This provides an empirical estimate of the effects of the additional restrictions involved – namely restrictions on the opening of hospitality venues and people meeting outdoors. We believe our study is also the first to investigate whether these effects differ by socioeconomic context.

### Implications of all the available evidence

Our study indicates that the introduction of Tier 3 restrictions to hospitality and meeting outdoors had a moderate effect reducing infections by 14% and 20% in October and December respectively, compared to the counterfactual of having introduced Tier 2 restrictions in those areas. We find no evidence that this impact varied by levels of socioeconomic deprivation. Although effect sizes were considerably higher in areas where there was a high prevalence of a new more infectious variant (B.1.1.7), this difference was not statistically significant at the 95% level – also potentially reflecting uncertainty in variant surveillance. This suggests that where higher levels of cases are clustered geographically a tiered response may be effective and is unlikely to increases inequalities in transmission.

## Methods

### Data and setting

We use data reported by the UK government on weekly number of people with at least one positive COVID-19 test result^15^ living in 7201 small areas in England called Middle Layer Super Output Areas (MSOAs), between September 2020 and January 2021. MSOAs are standard geographical units used to report statistics in England, with an average population of around 8000 people. In these data where there were fewer than 3 cases in any given week the number of cases was supressed to ensure individuals were not identifiable. In these situations, we imputed the number of cases, using complete data available at a higher geographical level (Local Authority), so that the sum of cases across MSOAs within a Local Authority (LA) was equal to the total number of cases reported for that LA in that week. In total, 4% of the outcome data was imputed in this way. LAs are municipalities covering the whole of England and have largely been the sub-national geographical units that have been used to organise response, testing and control measures during the pandemic.

As trends in reported cases will be affected by changes in testing capacity, testing strategy and public behaviour we adjusted the weekly count of cases reported for each MSOA by dividing it by a weekly estimate of the case detection rate in each LA area. To do this we compared the number of cases reported each day in each LA with the number of hospitalisations and deaths 8 days and 21 days later, respectively, using the method outlined by Kulu and Dorey.^16^ In brief we estimated the true weekly SARS-CoV-2 infection rate for each LA, using the age specific Infection Fatality Rate and Infection Hospitalisation Rate for the virus from Knock et al^17^ along with the daily number of deaths and hospitalisations for COVID-19 in each LA. This gave two estimates of infection rates, one from hospitalisation data and one from mortality date. We then took the average of these two estimates and calculated the case detection rate as the reported number of cases in each LA in each week divided by the estimated number of infections. The exact method is given in further detail in Appendix 5. In sensitivity analysis as outlined below we estimate all models without applying this case detection rate.

Alongside these data, we also measured local area characteristics that could potentially influence transmission and/or effectiveness of control measures. These included the English Indices of Multiple Deprivation (IMD) a composite measure of socioeconomic disadvantage^18^, average number of care home beds per head of population using data obtained from the Care Quality Commission, population density, the percentage of the population who were over 70 and the proportion of population aged 7-11 using 2019’s mid-year population estimates from the Office for National Statistics, the proportion of the population from Black Asian and Minority Ethnic (BAME) groups and the proportion of residents that were students, obtained from the 2011 Census. To additionally account for potential differences in testing between areas, we used data for LAs on the number of tests per head of population in the 4 weeks prior to the intervention available from the UK government COVID-19 dashboard. To account for differences in the prevalence of the more infectious variant of the virus, B.1.1.7^19^ that emerged in November we included data on the proportion positive tests with S-gene failure on PCR testing for each LA, using data from Public Health England.^20^

This time series of MSOA weekly data, area characteristics and linked LA data, was then merged with a dataset indicating the restrictions that each area was under in any given week. Data on restrictions was compiled and made available from the Open Data Intsitute.^21^

### Intervention

Tiered restrictions were introduced at two points in time. Firstly in October LAs in England, were placed in one of three Tiers with restrictions of increasing stringency.^22^ Tier 1 had the fewest restrictions, groups of up to 6 people were allowed to meet indoors or outdoors. In Tier 2, people were prohibited from mixing inside with individuals outside of their households, were only allowed to meet with up to 6 people outdoors, and pubs and restaurants had to close between 10pm and 5am. In Tier 3, people were additionally prohibited from meeting with people outside their household in private gardens. Pubs and restaurants were only allowed to remain open if they were acting as restaurants and serving a ‘substantial meal’. The Tier to which an area was allocated to was based on the average rate of change in case numbers and pressure on the health service across the LA.^23^ Although explicit criteria have not been published and there is some evidence of pressure from local politicians influencing decisions.^24^ Following this initial tiered system the whole of England was moved into a month-long national lock down from the 5th November before a return to a three-tier system at the beginning of December.^25^ This new tiered system had some differences in the restrictions for residents in each tier. The main difference was that in this second period in Tier 2 areas pubs and restaurants were only allowed to stay open if they were serving ‘substantial meals’, whilst in Tier 3 areas pubs and restaurants had to close except for providing takeaway food. The areas in each tier remained the same until 19^th^ of December, when some areas were re-assigned to a new ‘Tier 4’ on the 19^th^ December because of rapidly rising case numbers in certain areas, thought to largely be due to a new more infectious variant of the virus, B.1.1.7.^19^ In Tier 4 areas the guidance was to stay at home except for essential journeys. In both periods and in both Tier 2 and Tier 3, no mixing between households was permitted indoors.

### Analysis

For our analysis we investigate two intervention time points, weeks commencing 19^th^ of October and 7^th^ of December. In each period the initial allocation to tiers was announced on the Friday and we take the week starting the following Monday as the start of the intervention. We define MSOAs as being in the intervention group if they were in Tier 3 at those time points. We investigate the change in cases in the intervention group, four weeks before and after that time point, compared to a synthetic control group derived from places that entered Tier 2 at the same time. We analyse the two groups based on their initial allocation, even though in October 22% of the MSOAs initially allocated to Tier 2 were later moved to Tier 3. All the MSOAs initially allocated to Tier 3 in December stayed in Tier 3 until the country entered a national lockdown at the beginning of January 2021, apart from those in one local authority (Kent) which entered a new Tier 4 on the 20^th^ December. Analysing the groups based on their initial allocation is analogous to an intention to treat analysis in a trial and will provide a more conservative estimate of effect size. This will be less prone to bias that would result from selection of places based on their subsequent transition into Tier 3, which in itself would be influenced by the effectiveness of the tiered restrictions.^26^

To estimate the effect of Tier 3 restrictions compared to what would have been expected in those areas had they only been subject to Tier 2 restrictions we apply the synthetic control method for microdata developed by Robbins et al.^27,28^ The synthetic control method is a generalisation of difference-in-difference methods, whereby an untreated version of the treated cases (i.e., a synthetic control) is created using a weighted combination of untreated cases. Whilst previous synthetic control methods^29^ have been restricted to a single intervention unit often with limited data units available for the construction of a synthetic control, the method outlined by Robbins et al^27,28^ enables the use of multiple small areas in the intervention group and in constructing the synthetic control group. This offers several advantages as the large number of small-scale observations allows for better matching between treatment and synthetic control units. As the allocation to Tier 3 areas was based on the average level and trend in cases at the LA level this method is able to identify many small areas within Tier 2 areas that had similar levels and trends in cases as Tier 3 areas prior to the introduction of restrictions.

To construct the synthetic control group, we derive calibration weights to match the MSOAs in Tier 2 areas to Tier 3 areas across the 4 weeks prior to the intervention dates in terms of the local area characteristics described earlier and the prior four-week trend and levels in cases. For the October period we additionally included the number of weeks each area had been under local restrictions prior to the introduction of the tiered system. The weighting algorithm derives weights that meet a number of constraints. Firstly, the sum of weights in the control group equals the number of cases in the intervention group. Secondly, the weighted average of each of the local area characteristics in the synthetic control group matches those in the intervention group. Lastly, the synthetic control and intervention group also match across all pre-intervention time points in terms of the numbers of cases.^27^

The Average Treatment Effect for the Treated (ATT) is then estimated as the difference in cumulative number of cases in the intervention group (Tier 3) in the four weeks after the intervention time point, compared to the (weighted) number of cases in the synthetic control group. To estimate the 95% confidence intervals and p-values we apply a permutation procedure, through repeating the analysis through 250 placebo permutations randomly allocating Tier 2 MSOAs to the intervention group, to estimate the sampling distribution of the treatment effect and calculating permuted p-values and confidence intervals.^28^ All analysis was performed using R version 4.0.3 and the Microsynth package.^27^

To investigate whether there was a differential effect by socioeconomic group we grouped the MSOAs into 3 equal sized groups (tertiles) based on their level of deprivation as measured by the Indices of Multiple Deprivation. We then re-ran the weighting algorithm, stratifying the process by IMD tertile. As outlined by Robbins et al the Average Treatment Effect for the Treated (ATT) can be estimated from the data using the calibration weights in a weighted generalised linear model with the binary variable for the intervention group as the exposure. To investigate the differential effects by level of deprivation we fitted such a model with the stratified weights, but also included an interaction term between IMD tertile and the intervention indicator providing a measure of the intervention effect stratified by IMD tertile and a statistical test for an interaction effect. To account for the distribution of the outcome data and to estimate the relative effect we used a weighted Poisson model with a log link function.

As in December patterns of transmission and potentially the effectiveness of interventions were affected by the emergence of a new variant (B.1.1.7). We used the same approach as above for IMD to investigate differences in the effect of entering Tier 3 in December, based on the prevalence of the new variant in each area.

### Sensitivity analysis

As there was some uncertainty in the estimate of the case detection rate, we repeated the analysis without applying this adjustment. As there may be spill over effects with Tier 3 restrictions affecting and being affected by Tier 2 restrictions in neighbouring areas, we repeated the analysis excluding, from the synthetic control group, Tier 2 areas that were within 20 Km of Tier 3 areas. Our hypothesis was that the Tier 3 intervention effect could be mitigated or strengthened in those Tier 3 areas of neighbouring Tier 2 areas. It is possible that interventions in Tier 3 areas will affect neighbouring Tier 2 areas in either direction. For example, people might travel from Tier 3 areas to Tier 2 areas (against the official regulation) to partake in activities which are prohibited in Tier 3 areas, contributing to a rise in transmission in neighbouring Tier 2 areas. Alternatively, the implementation of Tier 3 intervention could reduce traffic flows and human interactions not only within the Tier 3 areas but also with neighbouring Tier 2 areas, which may benefit the neighbouring Tier 2 areas.

## Results

Figure 1 shows the areas that entered Tier 3 at each of the two time points. In October the initial Tier 3 areas (391 MSOAs) were entirely in the Merseyside and Lancashire areas in the North West of England. At the beginning of December, a larger proportion of England (2,858 MSOAs) initially entered Tier 3.

**Figure 1.**
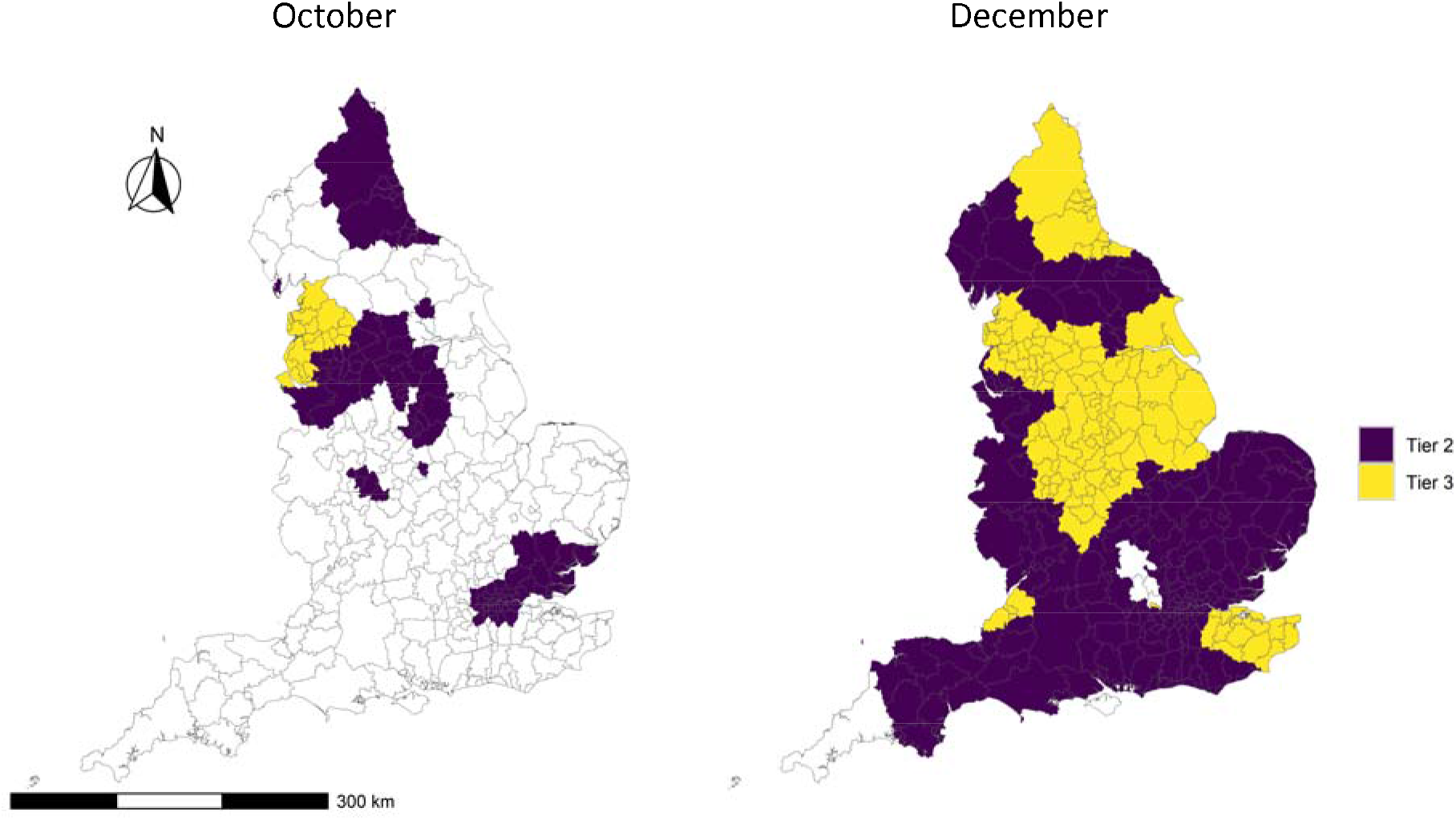
Location of areas that entered Tier 3 (yellow) and Tier 2 (purple) at the two intervention time points.

Table 1 presents summary statistics for areas within each Tier for both intervention time points. As would be expected estimated SARS-CoV-2 infection rates were higher in Tier 3 areas prior to the introduction of the tiered system at each time point. Tier 3 areas were more deprived on average at each time point. In both periods, Tier 3 areas tended to have a lower proportion of the population from BAME groups and lower population density. There were no differences in terms of students and care homes. In the December period the prevalence of the new variant was higher in Tier 2 areas compared to Tier 3 areas as measured by the proportion of S-gene target failure (SGTF) in routine PCR. In constructing the synthetic control group, weights were calculated to minimise the difference in each of the variables in Table 1. As an exact match was achieved in this weighting procedure the weighted average of each of these variables in the control group was identical to the intervention group. A map showing the geographical pattern of these weights is given in Appendix 2.

**Table 1.** The comparison between the Tier 3 and Tier 2 areas at the two intervention time points in the 4 weeks prior to the introduction of the Tiered system.

|  | October |  | December |  |
| --- | --- | --- | --- | --- |
|  | Tier 3 | Tier 2 | Tier 3 | Tier 2 |
| Average % estimated case detection rate (confirmed cases / estimated number of infections) in 4 weeks before Tiers introduced | 45 | 52 | 41 | 47 |
| Average tests per 100,000 per week in 4 weeks before Tiers introduced | 3232 | 2354 | 2970 | 2752 |
| Weekly infections per 100,000 per week in 4 weeks before Tiers introduced | 723 | 307 | 784 | 355 |
| Care home beds per 10,000 population | 98 | 69 | 86 | 80 |
| IMD score | 31 | 25 | 26 | 19 |
| Population density – people per hectare | 32 | 51 | 30 | 41 |
| % of population 70+ | 14 | 12 | 14 | 14 |
| % population 7-11 yrs old | 6 | 6 | 6 | 6 |
| % BAME | 7 | 22 | 12 | 15 |
| % *S-gene* target failure (SGTF) in routine PCR | NA | NA | 23 | 41 |
| % students | 3 | 4 | 3 | 3 |
| **Total population** | 3,068,261 | 25,272,230 | 23,347,218 | 31,682,197 |
| **Number of MSOAs** | 391 | 2994 | 2858 | 3774 |

Figure 2 shows the trend in infection rates in the intervention (Tier 3) and synthetic control areas before and after intervention at the two time points in October and December. As an exact match was achieved these trends are identical in the synthetic control group as in the intervention group in the pre-intervention period. In October the rates were increasing before the tiered system was introduced. They then started to fall, with the drop in cases starting first in the Tier 3 areas. With the second implementation of the tiered system in December infection rates were falling whilst the country was in national lockdown, then increased as the national lock down came to an end and the tiered system was introduced. This increase however was slower in the Tier 3 areas compared to the synthetic control. See Appendix 1 (Figure A and B) for charts displaying the differences between outcomes in the intervention and synthetic control groups compared to 250 placebo permutations.

**Figure 2.**
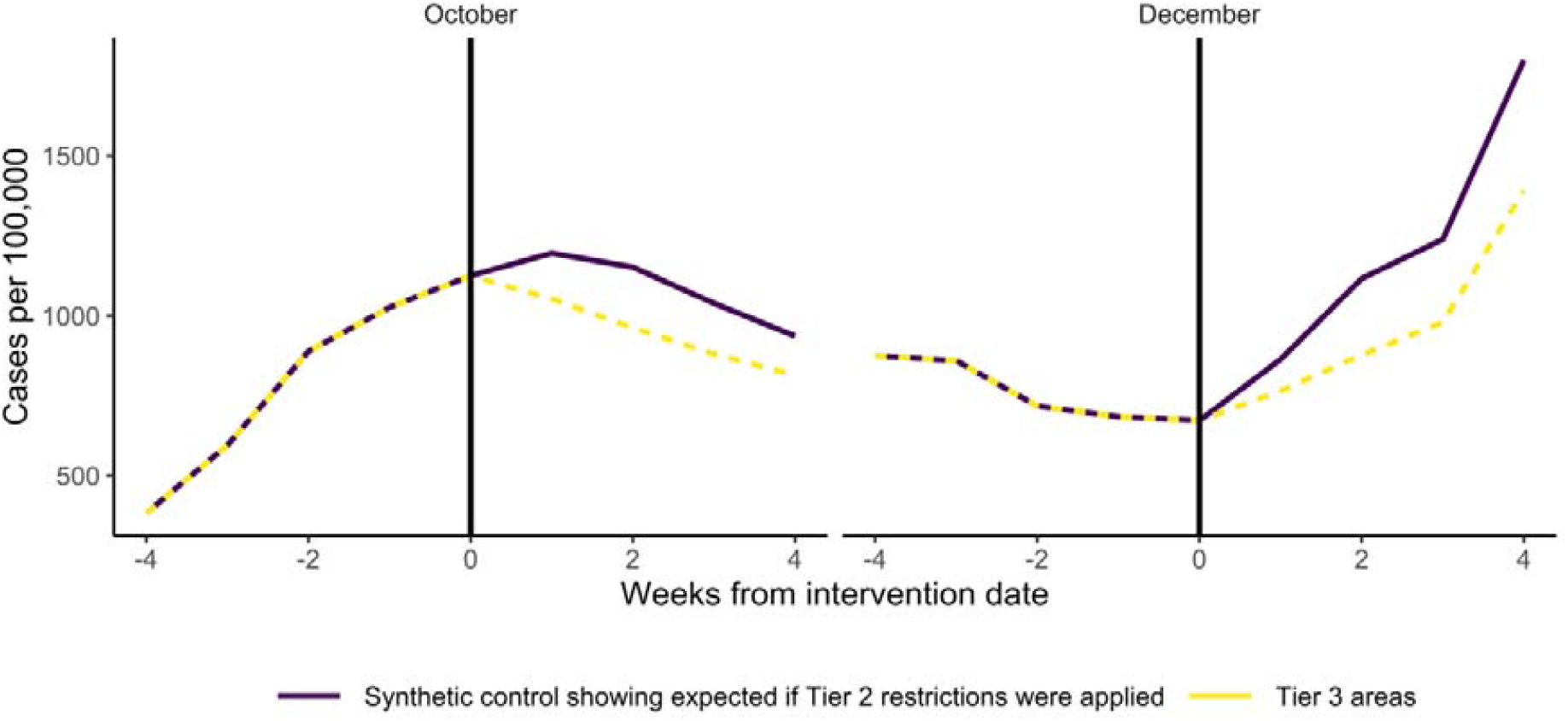
The trend in case rates in the Tier 3 areas and in the synthetic control group.

Table 2 presents the estimated effect, from the synthetic control analysis, of Tier 3 restrictions compared to what would have been expected if Tier 2 restrictions had been applied on those areas. In the October period the introduction of Tier 3 restrictions was estimated to have led to 14% fewer cases (95% CI 10% to 19%), than what would have been the case if Tier 2 restrictions would have been applied. In December, the Tier 3 restrictions are estimated to have led to a slightly greater reduction in cases, reducing cases by 20% (95% CI 13% to 29%).

**Table 2.** Results of the synthetic control analysis – indicating the relative reduction in infections in Tier 3 areas compared to what would have been expected if Tier 2 restrictions had been applied. Interaction analysis shows differences in effect by level of deprivation and prevalence of variant B.1.1.7 indicated by S-gene target failure (SGTF) where quantitative reverse transcriptase PCR is used for COVID-19 diagnosis.

|  | Percentage change in cases | 95% CI |  | p-value | p-value for interaction in subgroup analysis |
| --- | --- | --- | --- | --- | --- |
|  |  | LCL | UCL |  |  |
| **October - All Tier 3** | -14% | -19% | -10% | <0.001 |  |
| Most affluent areas | -10% | -17% | -2% | 0.016 |  |
| Intermediate deprivation | -15% | -22% | -7% | <0.001 | 0.354 |
| Most deprived areas | -19% | -29% | -7% | 0.003 | 0.214 |
| **December - All Tier 3** | -20% | -29% | -13% | <0.001 |  |
| Most affluent areas | -19% | -24% | -14% | <0.001 |  |
| Intermediate deprivation | -26% | -38% | -11% | 0.001 | 0.362 |
| Most deprived areas | -14% | -26% | 0% | 0.046 | 0.448 |
| Low SGTF (2%-20%) | -13% | -30% | 9% | 0.236 |  |
| Intermediate SGTF (21%-44%) | -6% | -16% | 5% | 0.271 | 0.579 |
| High SGTF (45%-85%) | -27% | -35% | -18% | <0.001 | 0.174 |

Through including an interaction term in the model to investigate differences in the effect across levels of deprivation, a statistically significant effect by each level of deprivation for each time period.This would suggest that Tier 3 interventions had an effect across all levels of deprivation. In October the effect estimate tended to be greater the more deprived an area was, although there was a high probability that this difference occurred by chance, with a p-value of 0.2 for a greater effect in the most deprived areas compared to the most affluent areas. In December, the effect was similar across all levels of deprivation and no interaction effects were detected again. Including an interaction term between the proportion of cases that were the new variant B.1.1.7 and the intervention group, suggested that the effect of Tier 3 restrictions may have been greater in areas where the new variant was more prevalent (−27%, 95% CIs –35% to –18%), but again the p-values for this interaction were greater than 0.05, indicating that these differences could have occurred by chance.

### Sensitivity analysis

To explore sensitivity to different assumptions, we used the confirmed COVID-19 cases instead of wider case-detection rates as our outcome and we found larger effects in both time periods (see Appendix 3). When excluding the Tier 2 MSOA areas located within 20 km of Tier 3 areas we found smaller effects but with high p values (see Appendix 4). This suggests that there may have been some spill over effects, whereby travel from Tier 3 areas to neighbouring Tier 2 areas contributed to a rise in transmission in neighbouring Tier 2 areas. However, such effects may well have occurred by chance.

## Discussion

Our study presents timely empirical evidence of the effectiveness of implementing regional tiered restrictions to manage COVID-19 responses. We find that more stringent regional restrictions can be effective at reducing infection rates. For both time periods, areas placed in Tier 3 restrictions experienced a moderate reduction in SARS-CoV-2 infections compared to what would have been expected if the same areas had been placed into Tier 2. This is consistent with reporting from the UK Scientific Advisory Group on Emergencies (SAGE) – which concluded in November that the October Tier 3 restrictions had reduced transmission, although they concluded that the effect could not be quantified and that it was unclear if Tier 3 restrictions alone would be sufficient to reduce R below 1.^30^ One simulation study predicted that moving into Tier 3 restrictions in England in October reduced the effective reproduction number (R) by 10%^3^ which is similar to the effect size we estimate here.

This study adds to the evidence that the additional restrictions on outdoor meeting and the hospitality sector have an important role to play in controlling transmission. The two main differences between Tier 3 and Tier 2 were that people in Tier 3 were not allowed to meet with people outside their household in private gardens and there were greater restrictions on pubs and restaurants. We cannot tell from our analysis whether either the restrictions on outdoor gathering or on hospitality settings, or a combination of both were responsible for the effects we observed. There is limited previous evidence, however, indicating that restricting outdoor contacts and hospitality reduces transmission, although it is thought these settings play a much smaller role than household transmission.^31,32^ Epidemiological analysis of outbreaks has noted that the largest super-spreading events have originated from pubs, clubs, restaurants, gyms and wedding venues.^33^ A report from the US found that those infected with SARS-CoV-2 without known close contact with confirmed cases were 3 times more likely to report dining at a restaurant than control participants^34^ and a systematic review has reported that pre-symptomatic transmission events frequently involve dining in close proximity.^35^ Outdoor proximity is thought to have a much lower risk of transmission than indoor proximity, with studies reporting a 19 times greater risk of transmission indoors compared to outdoors.^36^

Previous studies have not investigated whether tiered restrictions have different effect in different socioeconomic contexts. We find no consistent evidence that the effects differed by levels of deprivation. One might expect that some restrictions for example requirements to not to go to work, would have less of an effect on more disadvantaged groups in more precarious employment, where compensation is inadequate to enable them to stop working. The lack of differential effects that we see may be because the differences of restrictions in this study largely affect the hospitality sector and outdoor gathering and involvement in these activities may not differ between socioeconomic groups.

The emergence of new more infectious variants of SARS-CoV-2 raises concerns concerning the effectiveness of existing control measures. Increased infectiousness could mean that some activities such as outdoor contact previously seen as low risk could become higher risk. It is plausible that in the second period of our study, when the new variant B.1.1.7 was more prevalent that the marginal effect of restrictions on outdoor meeting and hospitality settings had a relatively greater effect on this more transmissible variant. We did find a larger effect in places with higher prevalence of the new variant – however there was high statistical uncertainty with this finding – possibly inflated by variant surveillance being more accurate in some areas than in others. Further investigation is needed to understand the effectiveness these restrictions in the presence of new variants.

A concern with localised restrictions in the control of COVID-19 is that there may be spill over effects, whereby, people move from areas of high restrictions to take advantage of lower restrictions (e.g. visiting restaurants and pubs) in neighbouring areas, increasing transmission in those areas and reducing the overall effectiveness of a system of differential local restrictions. We find some evidence for this, as when we excluded a 20 km buffer zone between Tier 3 and Tier 2 areas, the estimated effect of the Tier 3 restrictions was reduced. This may suggest that some of the increase in Tier 2, relative to Tier 3 areas was related to proximity between Tier 2 and Tier 3 areas.

Our analysis has some limitations. Firstly, we were only able to adjust for variation in the case detection rate using a relatively crude measure estimated at the LA level. This estimate assumes that the infection hospitalisation rate and the infection fatality rate does not vary between places that have similar prevalence of underlying health conditions and does not vary over the study time periods. Our analysis also assumes that the case detection rate is constant across MSOAs within each LA. We do however find larger effects when not applying our estimated case detection rate and we also adjusted for differences in the amount of testing carried out in each area. However differential changes in the case detection rate between intervention and control groups not accounted for in our estimates could bias our results. Secondly, although we were able to match areas to ensure a good balance of wider number of potential confounding factors prior to the intervention, there is still the potential that unmeasured variables could still contribute to biasing the results. Thirdly, we were only able to use data on small neighbourhood areas, rather than on individuals and therefore were unable to investigate how effects of control measures varied by individual or household characteristics – e.g. ethnicity, occupation or household size.

As countries such as the UK continue the battle to control COVID-19 cases, with large regional differences in transmission, tiered restrictions as well as national lockdowns will likely be needed to reduce infection levels whilst sufficient effective vaccination coverage is achieved. At present the UK government’s plans for exiting the current lock down are for a staged easing of restrictions at the same time across the whole of England^37^ with the easing of restrictions in outdoor contact occurring in the first stage (8 and 29^th^ March) and those related to indoor hospitality occurring during stage three (after 17^th^ May). Concerns have been raised about this – one size fits all – approach, given that high infection rates persist – particularly in more deprived areas.^38^ Our analysis indicates that tiered restrictions in outdoor gathering and in the hospitality sector are effective at moderately reducing the growth of cases and could be part of an effective strategy for reducing geographical differences in transmission risk as we emerge from the pandemic.

All the data we used are publicly accessible and statistical code is available from the authors on request.

BB, XZ are supported by the National Institute for Health Research (NIHR) Gastrointestinal Health Protection Research Unit. BB is also supported by the NIHR Applied Research Collaboration North West Coast (ARC NWC). GO is supported by the NIHR School for Public Health Research. IB is supported by NIHR Senior Investigator award. The views expressed in this publication are those of the author(s) and not necessarily those of the NIHR or the Department of Health and Social.

## Appendices

## Appendix 1. – Placebo permutations

Comparison of Tier 3 cases following introduction of tiered restrictions in October compared to a synthetic control.

**Figure A.**
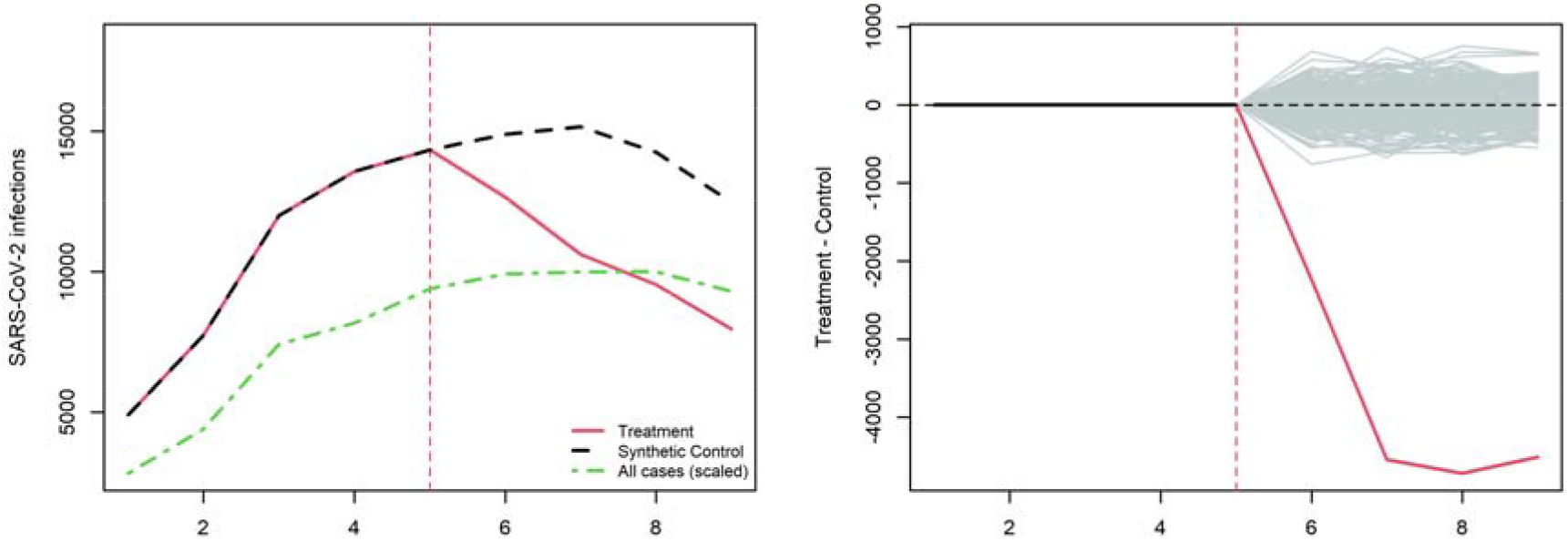
October all areas. Comparison of Tier 3 cases following introduction of tiered restrictions December compared to a synthetic control.

**Figure B.**
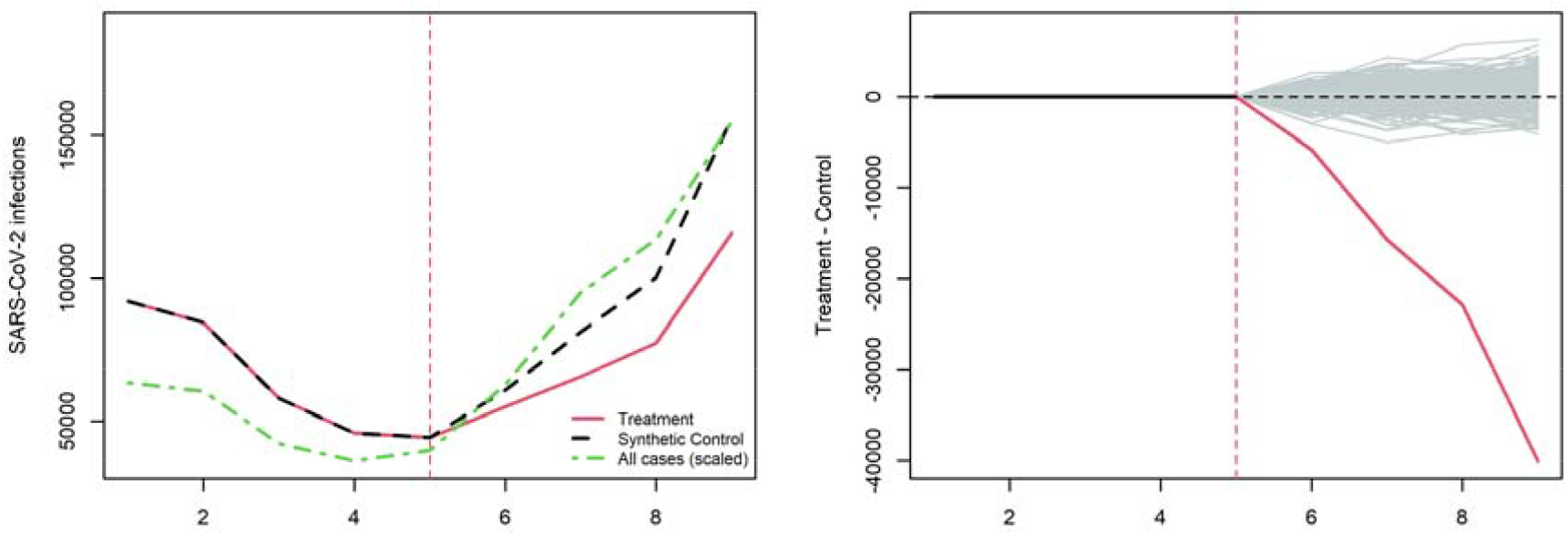
December all areas.

## Appendix 2. Weighting of Tier 2 areas used to construct synthetic control group

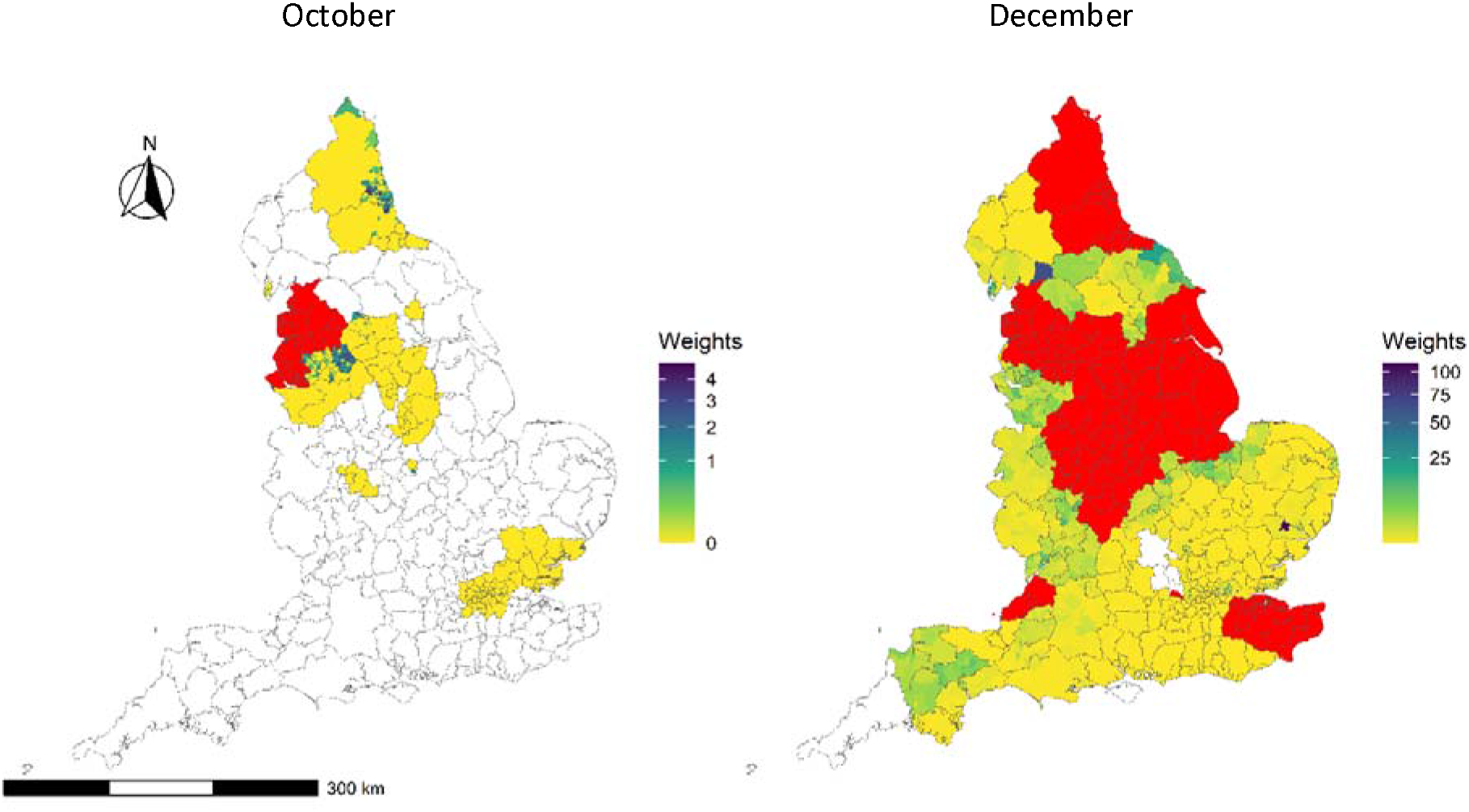

## Appendix 3. Replicating analysis only using confirmed cases as an outcome

**Table A.** The comparison of the number of confirmed cases between the Tier 3 and synthetic control Tier 2 areas during the two intervention periods, with lower (LCL) and upper (UCL) 95% confidence limits.

|  | Percentage change in cases | LCL | UCL | p-value |
| --- | --- | --- | --- | --- |
| **October 14<sup>th</sup> - All Tier 3** | -21% | -28% | -16% | <0.001 |
| **December 4<sup>th</sup> - All Tier 3** | -28% | -31% | -25% | <0.001 |

## Appendix 4. Sensitivity tests of the spatial spill over effect

In order to test the spatial spill over effect between the treatment (Tier 3) and comparison (Tier 2) group, we exclude the Tier 2 MSOA areas locating within 20 km of the treatment group. The distance between MSOA areas is measured by the Euclidean distance of the population weighted centroids of MSOA areas (Data source is ONS Geography Open Data https://geoportal.statistics.gov.uk/datasets/b0a6d8a3dc5d4718b3fd62c548d60f81_0).

**Table B.** The comparison of the number of cases between the Tier 3 and synthetic control Tier 2 areas during the two intervention periods, excluding the Tier 2 MSOA areas locating within 20 km of the treatment group.

|  | Percentage change in cases | LCL | UCL | p-value |
| --- | --- | --- | --- | --- |
| **October - All Tier 3** | -12% | -25% | 3% | 0.112 |
| **December - All Tier 3** | -12% | -28% | -2% | 0.128 |

**Figure H.**
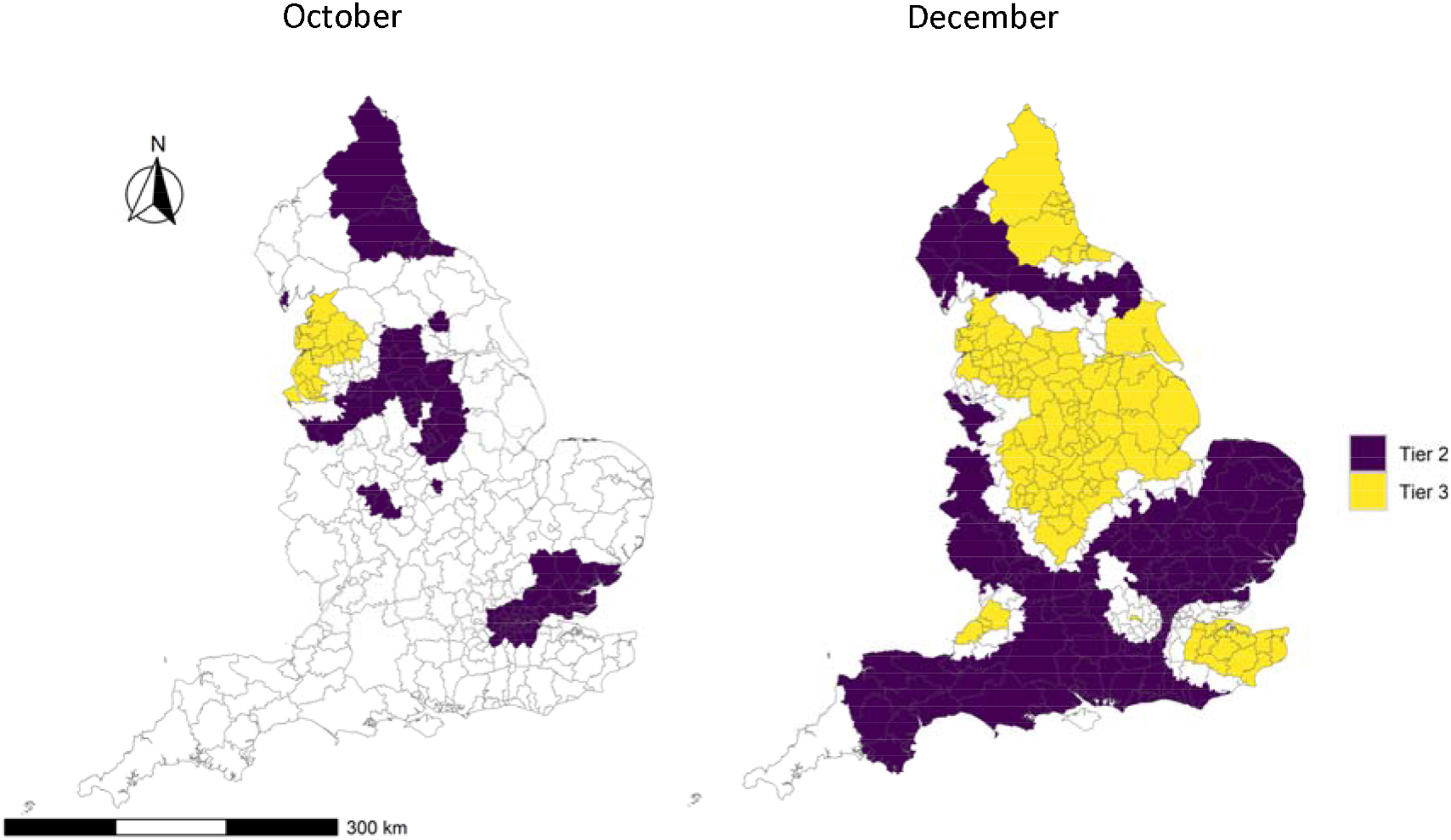
Location of areas that entered Tier 3 (yellow) and Tier 2 (purple) at the two intervention time points, excluding the Tier 2 MSOA areas locating within 20 km of the treatment group.

## Appendix 5. Calculating Local Authority level case detection rates

We use a method proposed by Kulu and Dorey to calculate local authority specific infection rates from data on observed and expected deaths and hospitalisations from Covid-19.^16^ The true infection rate was estimated from Covid-19 for each local authority as the number of observed deaths divided by the sum of the expected deaths for each age and sex sub-group, if everyone was infected. The expected deaths for each local authority was calculated by multiplying the population estimates for each age and sex group by the infection fatality rates for that sub-group. The same process was applied using hospitalisations instead of deaths. We use the Infection Fatality and Infection Hospitalisation Rates estimated by Knock et al.^17^

In order to account for varying rates of morbidity between local authorities, following Kulu and Dorey^16^ we apply a multiplier of chronic disease prevalence to the expected deaths and hospitalisation for each local authority. The idea being that in areas with higher rates of chronic disease, there would be expected to be higher COVID-19 Infection Fatality and Infection Hospitalisation Rates. Chronic disease was measured as the proportion of the population that had at least one admission to hospital with a diagnosis of cardiovascular disease, chronic respiratory disease, diabetes or chronic kidney disease recorded in their hospital record, which we found in previous research to be highly predictive of COVID-19 mortality.^5^ To calculate this multiplier we estimated the increased risk of COVID-19 mortality and hospitalisation associated with chronic illness for each local authority compared to the average using Poisson regression models.

The method outlined above was applied to 7 day moving averages of deaths and hospitalisation for each local authority. As hospital admissions and deaths occur sometime after initial infection, in order to estimate the true infection rate at a specific point in time we need to know the lag between initial case identification and both hospitalisation and death. To estimate this, we ran a set of regression models with observed hospital admissions or deaths as the dependent variable and with observed cases with a range of lags as the predictor variable. We tested lags of 1 to 20 days for hospitalisations and 7 to 30 days for deaths with each different lag tested in a separate model. We determined the best fit lag by selecting the model with the lowest Akaike Information Criterion (AIC). This was 8 days for hospitalisations and 15 days for deaths.

We then divided the 7 day’s rolling average of observed cases for each day by the estimated number infected calculated separately using hospitalisation data and deaths data and then took the average of the output of the two methods as our measure of case detection rate. Where there was missing data for either hospitalisations or deaths we took the estimated number infected using the method where there was complete data.

